# Ambient AI Documentation in Clinical Genetics: Perspectives on Implementation and Impact on Burnout

**DOI:** 10.64898/2026.06.30.26356723

**Authors:** Anjali Narain, Jason Misurac, Jennifer Van Tiem, Chase LaSpisa, Colleen A. Campbell

## Abstract

**Objectives:** To assess genetic counselors’ perspectives on ambient AI adoption and its impact on counselor burnout.

**Materials and Methods:** We utilized a mixed methods approach, surveying burnout using the validated Stanford Professional Fulfilment Index (PFI) before and after ambient AI adoption and exploring adoption perspectives through semi-structured interviews.

**Results:** 64% of participants (16/25) completed the pre-survey, with eleven completing post-surveys (69% response rate for completion of all three surveys). 14/25 participants completed interviews. Ambient AI use was associated with reduction in burnout after 90 days; respondents who reported using ambient AI (vs. non-use) had burnout scores 1.05 points lower, on average (p=0.008). Benefits of adoption included effective use with interpreters, memory aid, summarization of non-templated note sections (e.g. family/social history), and improved patient engagement. Challenges included template customization, variable accuracy, oversimplified medical language, and rapport disruption during consent. Ethical and regulatory considerations included data privacy, bias, awareness of training resources, and concerns about job displacement.

**Discussion:** Ambient AI documentation can reduce documentation burden and burnout among genetic counselors. By evaluating both outcomes and real-world implementation considerations, our study provides evidence to guide scalable integration of AI-enabled documentation tools in clinical genomic medicine.

**Conclusion:** Ambient AI can help support the sustainability of the clinical genetics workforce as genomic medicine initiatives are scaled across health systems. Addressing genetics-specific documentation needs while prioritizing patient trust, transparency, and provider oversight is essential for responsible ambient AI implementation.

## 1. BACKGROUND AND SIGNIFICANCE

Ambient artificial intelligence (AI) documentation is increasingly being adopted across health systems to streamline clinical encounters, with 63% of Epic centers implementing ambient documentation.^1^ Ambient AI tools transcribe patient-provider conversations during a visit and use generative AI to draft a clinic note for review and editing, functioning as computerized scribes. Prior studies examining the implementation of ambient AI across multiple specialties have shown reductions in provider burnout and documentation burden and improvements in patient engagement.^2–11^ However, the applicability and performance of these tools can vary across specialties, and the role of ambient AI in supporting genomic medicine workflows remains incompletely characterized.^7^

While the demand for clinical genetics services continues to expand with rising population health screening initiatives and the increasing utility of exome and genome sequencing, limited access to genetics professionals and inefficient systemic workflows, including documentation, limit health systems’ ability to meet this demand.^12^ Genetic counselors play a key role in the delivery of genomic medicine, yet 64% of their time is spent on non-patient-facing activities including documentation thereby limiting their ability to meet this demand.^13^ Together, these challenges contribute to high rates of burnout in the clinical genomics workforce.^14–17^ The downstream impacts of burnout include genetics professionals leaving the field and decreased quality of patient care in genetics visits through reduced counseling effectiveness, empathy, and positive regard.>^14–15, 18–21^

Since ambient AI use has been associated with reductions in burnout for other healthcare providers, its implementation in clinical genetics could be a promising intervention to reduce documentation burden and help scale genomic health programs as the demand for clinical genetics services continues to increase.^2–12^ Genetic counseling documentation often includes concise descriptions of rare genetic conditions, extensive psychosocial counseling, and ethical considerations, highlighting the need to evaluate ambient AI tools within this distinct clinical context. No studies thus far have explored the adoption of ambient AI tools for genetic counseling documentation. As early adopters of an ambient AI documentation tool integrated into the electronic health record (EHR) at the University of Iowa (UI) Health Care, and as an institution with a large genetic counseling service distributed across multiple specialties, we are well-positioned to address this gap.

## 2. OBJECTIVES

The first objective of our study is to evaluate the effect of ambient AI on genetic counselor burnout. Our second objective is to explore counselor perspectives to identify barriers and facilitators influencing adoption of ambient AI in genomic medicine workflows.

## 3. MATERIALS AND METHODS

The University of Iowa Institutional Review Board (IRB) determined that this project did not constitute human subjects research (IRB #202508020). Participants were informed about the purpose of the study, and participation in surveys and interviews was voluntary. Completion of surveys implied consent. For interviews, individuals who agreed to participate following email invitation and proceeded with the interview were considered to have provided consent.

This was a mixed methods study, combining pre-post observational surveys to measure genetic counselor burnout with qualitative semi-structured interviews to explore perspectives on ambient AI documentation. CORE-Q Guidelines were implemented for the qualitative arm of this project.^22^

### Study Population and Recruitment

The study population comprised 25 genetic counselors employed at UI Health Care across nine different specialties (Supplemental Table 1). Convenience sampling was used to recruit participants via email and face-to-face interactions to complete burnout surveys and participate in semi-structured interviews from August 2024 - December 2025.^23–24^

### Data Collection

#### Quantitative Arm

Burnout was assessed using the Stanford Professional Fulfillment Index (PFI) survey, a validated burnout instrument that measures professional fulfillment, work exhaustion, and interpersonal disengagement.^25^ The study population was recruited to complete PFI surveys via Qualtrics at three timepoints: pre-ambient AI (prior to ambient AI’s integration into the EHR), 90 days after pre-survey completion, and 270 days after pre-survey completion. The pre-ambient AI and 90-day surveys were distributed to genetic counselors by UI Health Care as part of a hospital-wide project evaluating ambient AI for physicians and APPs, and these responses were obtained.^26^ The 270-day survey mirrored the 90-day survey. All surveys included open-ended comment boxes. These timepoints were selected to capture both short-term and longer-term changes in burnout levels associated with ambient AI usage.

#### Qualitative Arm

To assess genetic counselor perspectives, a genetic counselor, AN, with experience in qualitative research conducted virtual semi-structured interviews between September and December 2025. The interview guide was adapted by AN from a previous guide used by an implementation scientist, JVT, in interviews with physicians and advanced practice providers (APPs) at UI Health Care on the same topic.^27^ Participants were invited to participate regardless of whether they had used the ambient AI tool and were informed of the project goals. Data saturation was defined as when no new information or ideas emerged from following interviews.^28^

### Data Analysis

#### Quantitative Arm

Burnout scores were analyzed by comparing pre- and post-survey PFI scores across the three timepoints using two statistical tests: 1) Linear mixed-effects model and 2) related-samples Wilcoxon signed-rank test.

To examine changes in burnout following access to ambient AI, we estimated a linear mixed-effects model with random intercepts for individuals to account for repeated observations and unobserved individual heterogeneity. Models were estimated using maximum likelihood and retained all available observations.^29^ ‘Wave_post’ is a continuous time indicator for post-baseline measurement occasions, coded 0 for the 90-day survey and 1 for the 270-day survey. It captures elapsed time since the first post-exposure measurement, allowing burnout trajectories to be modeled linearly after ambient AI access. ‘Wave_post x Ambient AI use’ is an interaction term that tests whether the association between ambient AI usage and burnout differs across post-baseline survey waves. A positive coefficient indicates that burnout increased more among users relative to non-users. A non-significant coefficient suggests similar post-exposure burnout trajectories across groups. A detailed description of this analysis is in the Supplemental Methods section.

To complement these analyses, we used the Wilcoxon signed-rank test to assess paired differences in burnout scores across baseline, 90-day, and 270-day measurements, stratified by ambient AI use. This non-parametric approach avoids assumptions of normality and evaluates both magnitude and direction of change.^30^

#### Qualitative Arm

Interviews were audio-recorded and transcribed, and transcripts were accuracy-checked. AN and JVT generated a codebook using inductive and deducting coding.^31^ An initial codebook, developed for prior research on physician and advanced practice provider perspectives on ambient AI at UI Health Care, was applied to the first transcript by AN and JVT.^27^ Inductive codes were added to capture themes specific to genetic counseling. Coding was performed using MAXQDA, with AN coding all transcripts and JVT co-coding every 3 transcripts to ensure consistency.^32^ Thematic analysis was performed by AN and the Consolidated Framework for Implementation Research (CFIR) model was used as a framework for this analysis.^33–34^

## 4. RESULTS

Demographics of participants who completed surveys and interviews are depicted in Supplemental Table 1. Eight participants completed all three burnout surveys and interviews.

### A. QUANTITATIVE BURNOUT ANALYSIS

16 out of 25 invited participants completed the pre-ambient AI survey (64% response rate). 14 of these respondents were female (87.5%), 12 in the 25-34 years age range (75%), and 10 had 0-5 years of work experience working at UI Health Care. Out of 16 pre-survey respondents, 11 completed the subsequent 90-day and 270-day surveys (69%). Supplemental information regarding response rates is outlined in Supplemental Table 2.

Table 1 presents the results of the linear mixed-effects model estimated using Wave_post as a fixed effect and a random intercept for survey respondents to determine changes in burnout over time. The estimated average burnout score at the 90-day survey timepoint for respondents who reported not using ambient AI was 3.5 (intercept). At the 90-day survey timepoint, respondents who reported using ambient AI (vs. non-use) had burnout scores about 1.05 points lower, on average (p-value=0.008). Among reported non-users, burnout changed very little between 90 and 270 days and is not statistically significant (coefficient of 0.023, p-value=0.907). As for the interaction, the change in burnout between 90 and 270 days does not significantly differ between ambient AI users and non-users. Ambient AI users’ burnout may have increased slightly faster (positive coefficient), but uncertainty is large due to small sample sizes.

**Table 1:** Linear Mixed-Effects Regression Model for Burnout Score Across Survey Wave.

| Variable | Coefficient | Std. Error | z-score | p-value | 95% CI |
| --- | --- | --- | --- | --- | --- |
| **Fixed effects** |  |  |  |  |  |
| Wave_post (time since pre-survey completion) | 0.023 | 0.195 | 0.12 | 0.907 | [−0.359, 0.405] |
| Ambient AI use | −1.050 | 0.396 | −2.65 | 0.008 | [−1.826, −0.273] |
| Wave_post × Ambient AI use | 0.353 | 0.463 | 0.76 | 0.445 | [−0.554, 1.260] |
| Intercept | 3.508 | 0.455 | 7.71 | <0.001 | [2.616, 4.399] |
| **Random effects** |  |  |  |  |  |
| Variance (Intercept \| Respondent) | 2.631 | 1.055 |  |  | [1.199, 5.775] |
| Residual variance | 0.553 | 0.155 |  |  | [0.320, 0.958] |
| **Model fit** |  |  |  |  |  |
| Observations (N) | 41 |  |  |  |  |
| Respondents | 15 |  |  |  |  |
| Wald $\chi^2$ (df = 3) | 8.57 | | | 0.036 | |
| Log likelihood | −65.74 |  |  |  |  |
| LR test vs. linear model | $\chi^2 = 28.03$ | | | <0.001 | |

Figure 1a depicts participants’ burnout scores at the pre-survey and 90-day timepoints stratified by ambient AI users vs non-users, with Figure 1b adding the burnout score distributions at the 270-day timepoint. In Figure 1b, n is the compiled number of survey respondents reporting Ambient AI use versus non-use across timepoints, with the same 15 survey respondents. The validated cutoff for overall median burnout using the Stanford PFI is 3.325 on a 10-point scale.^25^ To further analyze differences in median burnout scores across time and ambient AI use, a series of Wilcoxon sign-ranked tests were conducted.

**Figure 1:**
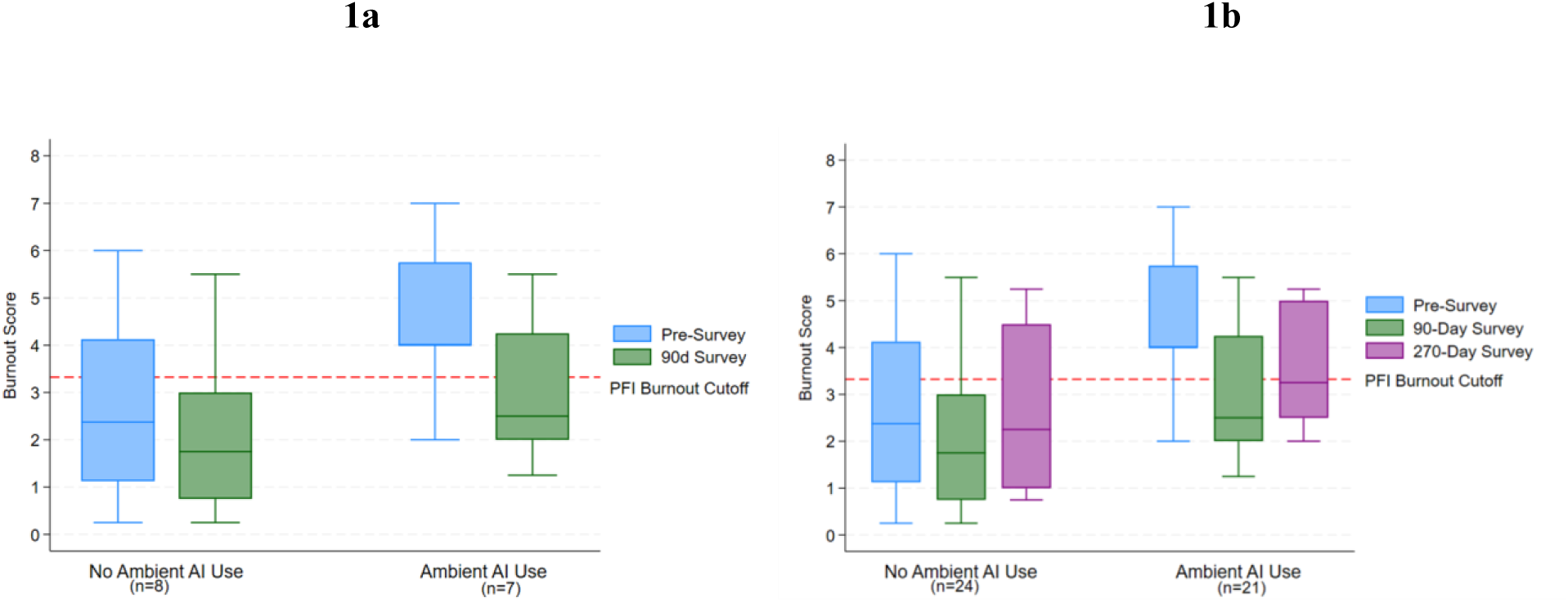
Box Plot of Burnout Scores by Ambient AI Usage Across Time (with PFI burnout cutoff of 3.325).

The median burnout score for participants who reported ambient AI use decreased from 4 at the pre-survey mark to 2.5 at the 90-day mark (Figure 1b). The Wilcoxon sign-ranked test indicated that there is evidence of a statistically significant negative difference from zero in median scores for reported ambient AI users from the pre-survey to the 90-day survey level (z = 2.31, exact p = 0.031; n = 7). The median score for ambient AI users increased slightly from the 90-day survey level to about 3.25 at 270-day mark (Figure 1b), but the Wilcoxon test did not indicate a statistically significant difference from zero in the medians. For non-users, the median burnout score decreased from 2.4 at the pre-survey mark to 1.75 at the 90-day mark and was 2.25 at the 270-day mark. The Wilcoxon sign-ranked tests did not indicate that the median differences significantly differ from zero for non-users at any point from baseline to the 270-day mark.

These figures thus indicate very little change in the distribution and median burnout scores over time for non-users, but ambient AI users saw a reduction in burnout scores compared to non-users. Burnout scores for ambient AI users first decreased at the 90-day mark, and then slightly increased at the 270-day level, though the change was not statistically significant. This indicates a potential short-term influence of ambient AI usage on burnout. The linear mixed effects model results indicate that survey respondents using ambient AI had burnout scores around 1.05 points lower, on average (p-value= 0.008).

Figure 2a displays the proportion of survey respondents experiencing burnout at the pre-survey and 90-day survey timepoints, stratified by reported ambient AI usage vs non-usage at the 90-day mark (extrapolated again to the pre-survey level). The validated PFI burnout cutoff of 3.325 was used to determine whether respondents were experiencing burnout. Among ambient AI users, the proportion of participants experiencing burnout decreased from 86% at the pre-survey timepoint to 43% at 90 days. For non-users, the proportion of participants experiencing burnout remained constant at 38%. When including burnout at the 270-day timepoint (Figure 2b), the proportion of ambient AI users experiencing burnout stayed constant at 43% across 90 days and 270 days. The proportion of non-users experiencing burnout was 38% at 90 days and 50% at 270 days. While not calculated statistically, we observe that at the pre-survey timepoint, the number of users experiencing burnout was higher than non-users, making it possible that those who were more burned out at the timepoint were more motivated to use ambient AI.

**Figure 2:**
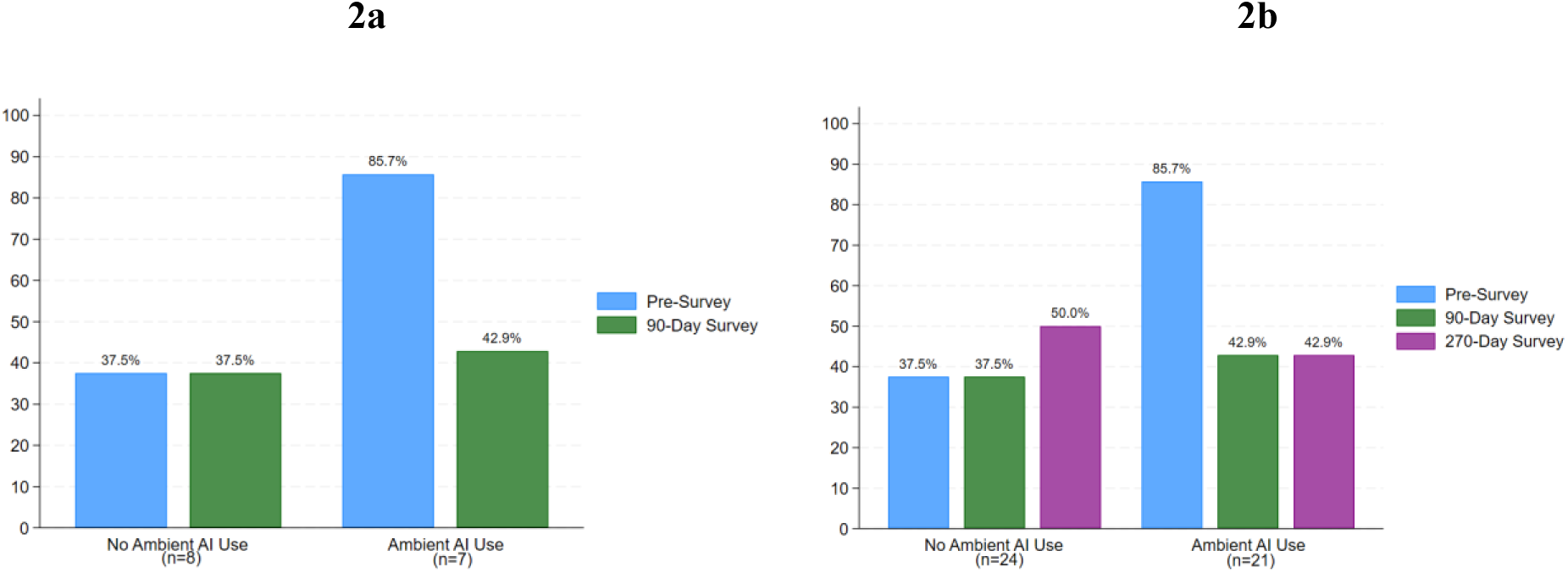
Proportion (percentage) of survey respondents with burnout (PFI cutoff of 3.325) across time and by ambient AI.

#### Descriptive Statistics

At the pre-survey timepoint, 75% (12/16) of genetic counselors stated they spent more than 5 hours per week on documentation. After using ambient AI, the median perceived time savings per week was 1.1 hours at the 90-day timepoint (n=6) and 2 hours at the 270-day timepoint (n=7). Participants most often used the ‘Custom Instructions’ feature, a tool that allows users to customize different sections of their note template according to their preferences. The most important driver for edits was missing information or notes not being in the user’s style. 100% of users agreed (somewhat agreed or strongly agreed) that ambient AI made them less worried about forgetting elements of the patient visit. 71% (5/7) users stated they would like educational materials/tipsheets on how to customize this tool emailed to them, one stated they’d like to set up a 1-on-1 meeting for ambient AI user support, and four stated they would like a presentation about the ambient AI scribe for their department.

### B. QUALITATIVE INTERVIEW ANALYSIS

14 out of 25 study participants completed interviews lasting an average of 21 minutes (ranging 12 minutes - 51 minutes).^35^ Data saturation was reached by interview 13.^28^

Thematic analysis revealed five themes: 1) Adoption and usage 2) Benefits 3) Challenges 4) Accuracy 5) Ethical, Legal, and Social Issues (ELSI). Quotes supporting these themes and subthemes are listed in Table 2.

**Table 2:**
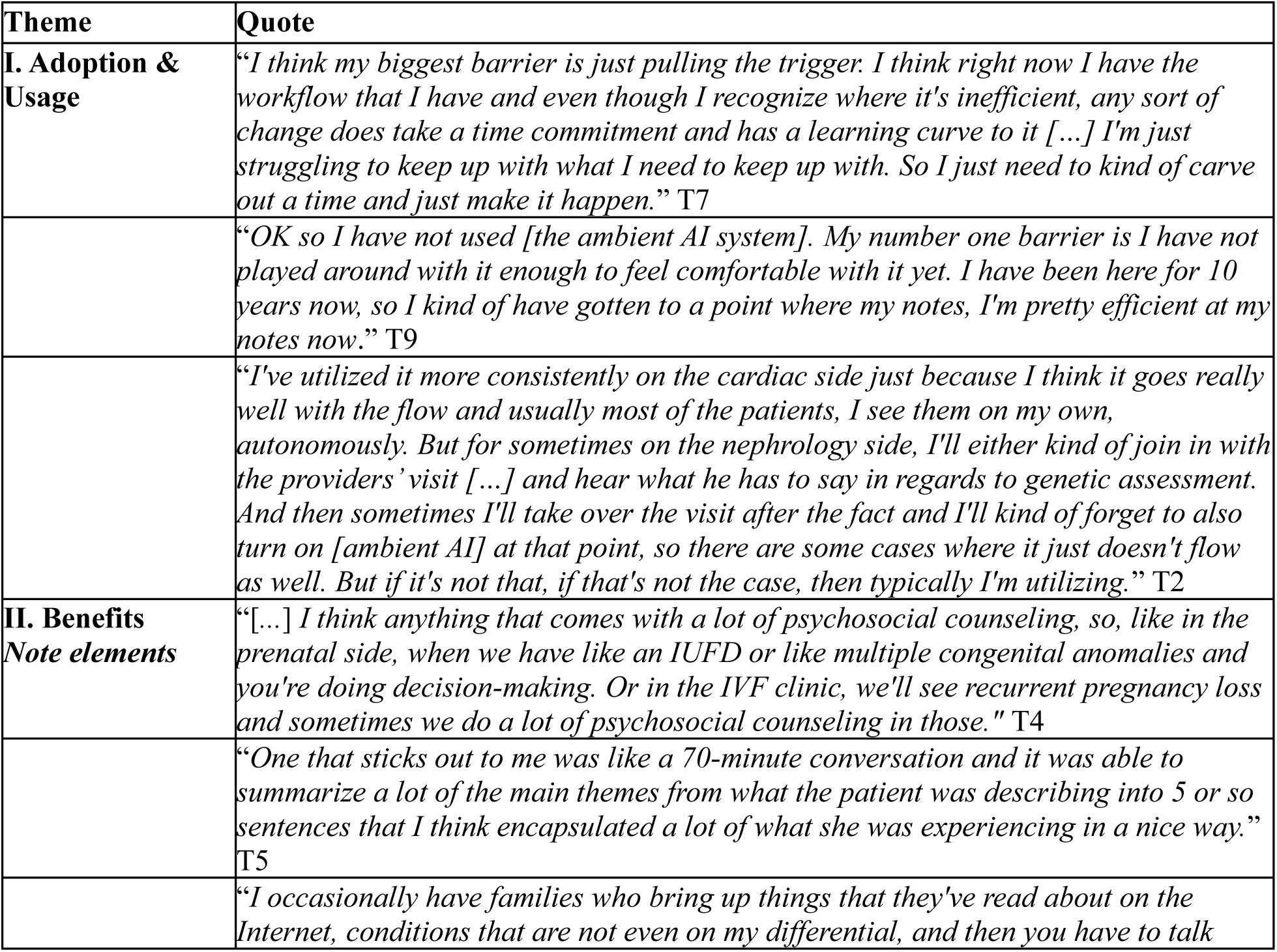
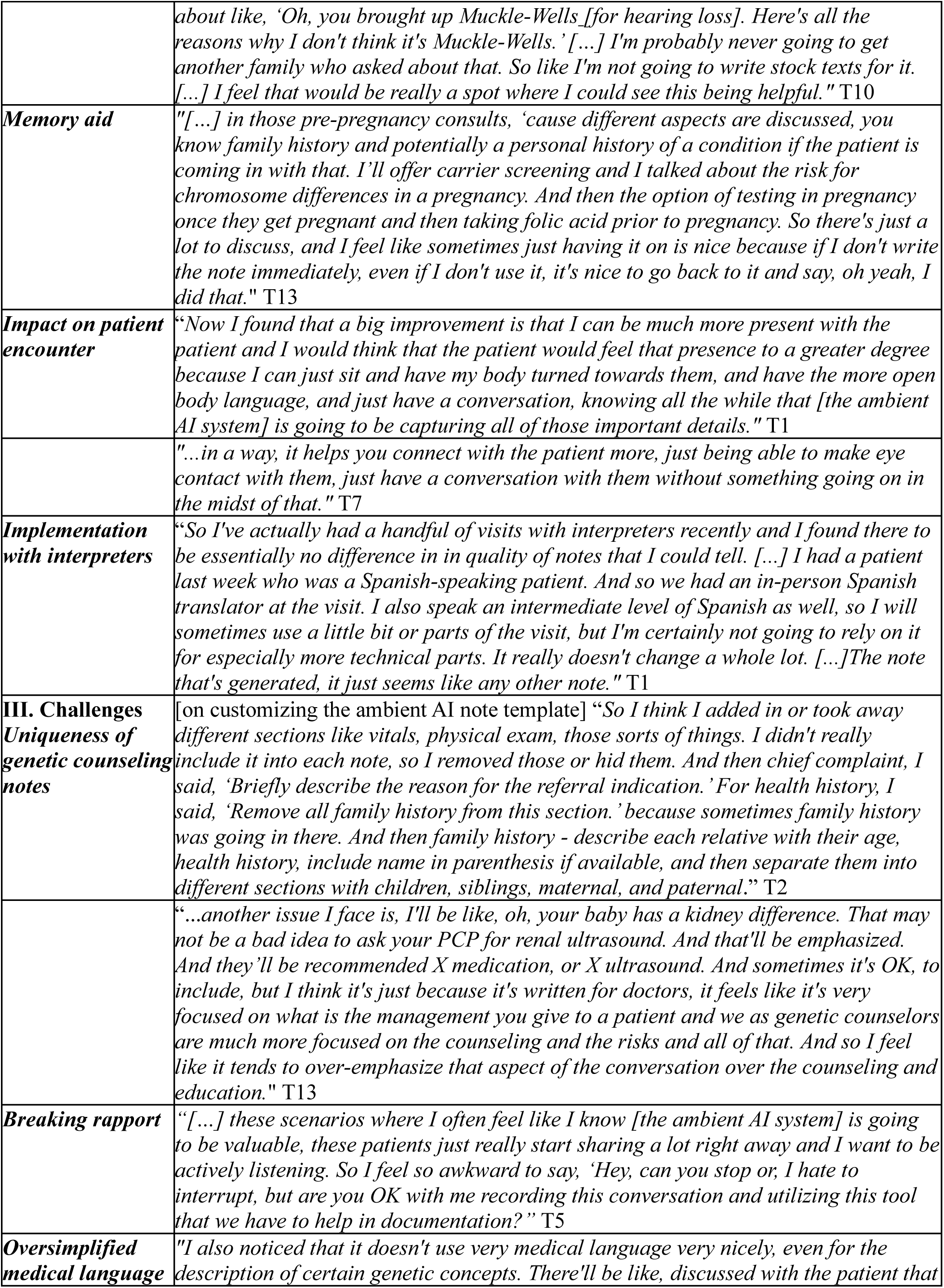
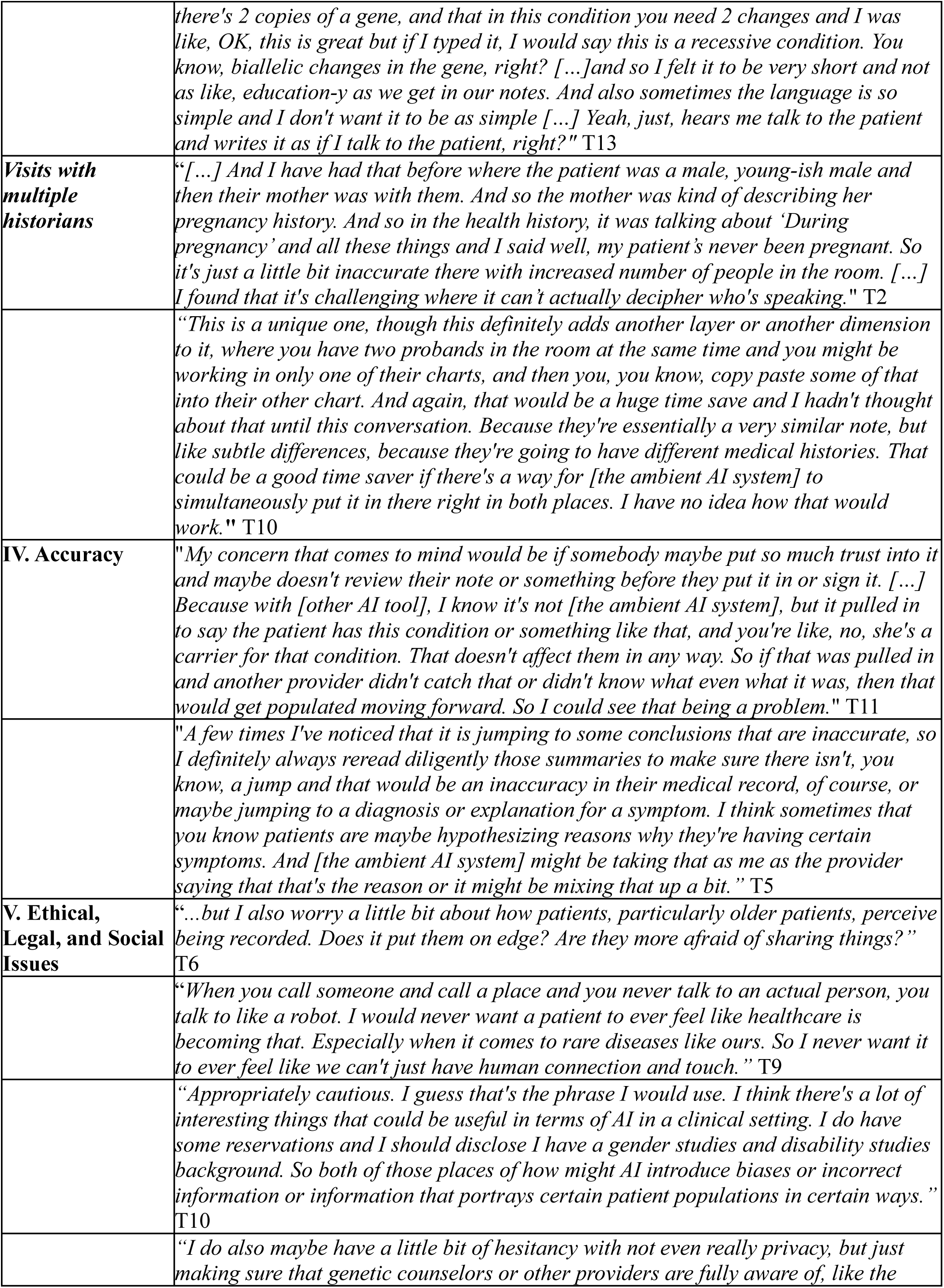
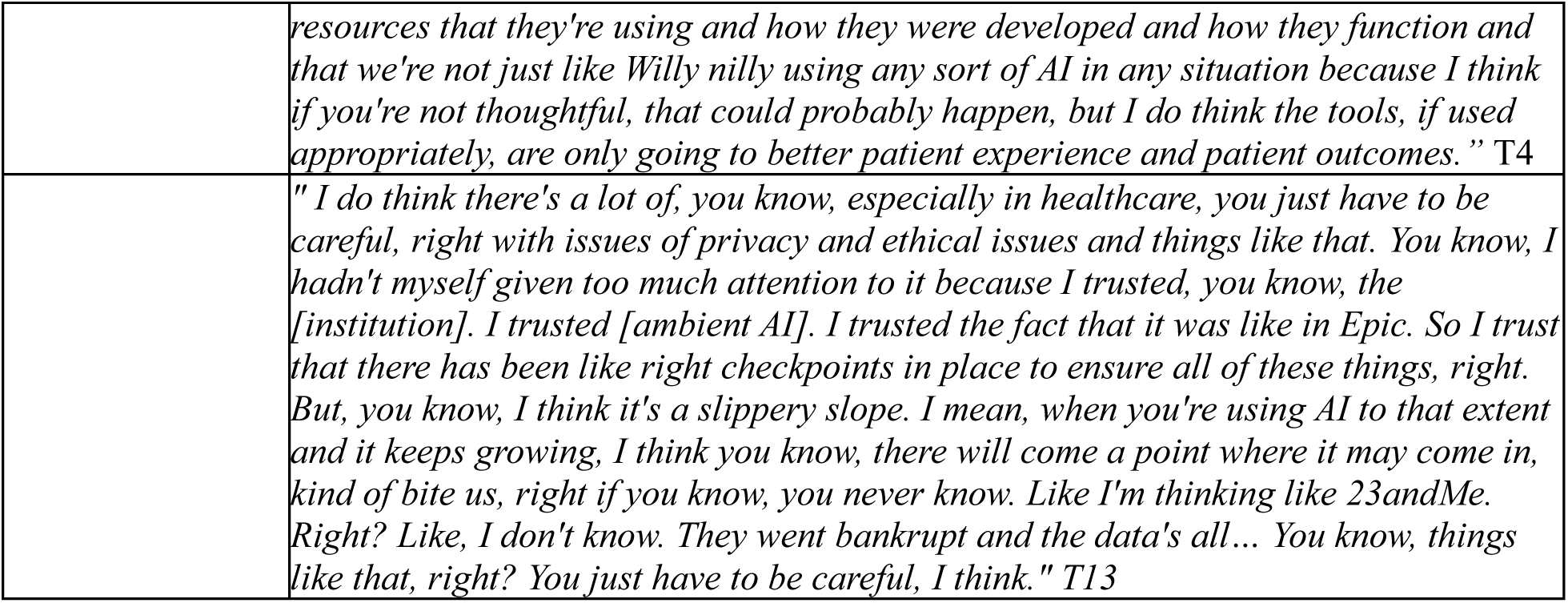
Quotes supporting themes and subthemes from thematic analysis.

#### I. Adoption and Usage

Participants were a spectrum of early to late or non-adopters of ambient AI. Seven participants had adopted ambient AI at the time of the interview, while seven had not adopted the tool. Several factors influenced non-adoption. Participants who were satisfied with their current documentation workflow and had sufficient dotphrases for their notes were not motivated to adopt this tool. Among those who were dissatisfied with their current workflow and described it to be inefficient, lack of time or not being tech-savvy to navigate the learning curve or experiment with the tool were barriers to adoption. Forgetting to turn on this tool was another barrier for both adoption and usage.

Usage varied among adopters. High users were defined as though who had used ambient AI for almost every visit, moderate users had used ambient AI often or for certain visit types but not consistently for every visit, and low users had used ambient AI sporadically or had experimented with it a few times. Two participants were high users, two were moderate users, and three were low users. Frequency of usage was influenced by visit type (genetic counseling-only visit or group visit with a doctor), with some stating that ambient AI didn’t go with the flow of a visit with multiple providers. Many reported increased use to summarize conversations for which dotphrases did not exist, such as sessions that were more focused on psychosocial counseling. One participant’s usage rose when they had more time to personalize the tool and fell when they were overwhelmed with day-to-day tasks.

When asked about general adoption of new tools or technologies, middle or late adopters stated reluctance to be early adopters as they wanted to avoid troubleshooting early technical issues and preferred adoption after initial issues had been resolved.

#### II. Benefits of ambient AI

##### a. Note elements

All ambient AI users reported selectively copying certain sections of the AI-generated draft note rather than the entire note. Many valued the tool’s ability to summarize non-templated note elements including family history, psychosocial conversations, decision-making discussions (such as factors influencing a patient’s choice to pursue genetic testing and select an appropriate test), rare genetic conditions, and non-templated telephone notes. Some reported benefit in summarizing the medical history/HPI and plan. In many cases, counselors noted that comprehensively summarizing these sections required additional prompting.

For note sections that could be summarized by a dotphrase, participants preferred dotphrases over AI-generated text. Dotphrases had standardized language and formatting, thus ensuring consistency across patients especially when summarizing the description of common genetic conditions or the types of genetic test results.

##### b. Memory aid

A majority of ambient AI users found it useful as a memory aid, expressing relief in being able to review the session transcript to either catch forgotten details or cross-check collected information for accuracy.

##### c. Impact on patient encounter

Ambient AI users could focus less on typing, thus better engaging with patients through increased eye contact and improved body language.

##### d. Implementation with interpreters

Participants who had used ambient AI with an interpreter present during the appointment did not encounter any challenges with note generation and reported no difference in the quality of the generated note.

#### III. Challenges

##### a. Uniqueness of genetic counseling notes

Since current ambient AI note templates are designed primarily for documentation needs of physicians, advanced practice providers (APPs), and nurses, and are primarily in the ‘Subjective, Objective, Assessment and Plan’ (SOAP) format, counselors found that ambient AI overemphasized SOAP-related note sections while under-emphasizing genetic counseling note sections. They thus had to customize their note templates to ensure the necessary genetic counseling note components were included. Customization strategies included adjusting formatting requirements, removing irrelevant SOAP sections (vitals or physical exam), and using ambient AI’s features such as ‘Custom Instructions’ (custom prompt instructions applied to each subsequent note) or ‘Magic Edit’ (regenerate a specific note with additional instructions) to add required note elements. However, many reported lacking the time or technical proficiency to make these modifications.

##### b. Breaking rapport

Some reported not using ambient AI when sessions began with heavy psychosocial counseling. Given the sensitive nature of such sessions, participants expressed concern that interrupting such a conversation to obtain consent for ambient AI use and informing them that the conversation would be transcribed verbatim would break rapport.

##### c. Oversimplification of medical language

A few participants expressed that the ambient AI-generated paragraphs lacked precise medical language, with genetic concepts such as inheritance patterns being oversimplified. Since clinic notes are often reviewed by other providers, participants emphasized the need for accurate medical terminology.

##### d. Visits with multiple historians

Participants implemented ambient AI in group settings in two scenarios. The first involved a proband accompanied by a caregiver or family member who contributed to the family history. In this scenario, attribution errors occasionally occurred, with ambient AI populating a family member’s history (typically entered in the family history section) as part of the proband’s medical history. In the second scenario, when two probands were present during the same visit, ambient AI was unable to populate two medical records simultaneously.

#### IV. Accuracy

Participants had mixed opinions on the accuracy of generated notes. While many appreciated the transcript serving as a memory aid for improving note accuracy and comprehensiveness, some expressed worry about inaccuracies in generated content. These participants stated the need for corroboration of AI-generated notes to prevent communication errors among providers and prevent propagation of inaccuracies into future documentation by other providers.

Additionally, although ambient AI lacks reasoning capabilities and functions only by summarizing the transcript, one participant experienced over-inference and perceived ambient AI to jump to inaccurate conclusions about a patient’s diagnosis.

#### V. Ethical, Legal, and Social Issues (ELSI)

While participants were overall supportive of integration of AI tools into clinical workflows, they expressed that it should be used thoughtfully. Several concerns were raised, with the most prominent one being patient and provider lack of trust due to data privacy and security concerns. This concern was stated despite the institutional policy that providers obtain consent from patients before using ambient AI in a clinical visit. Additional concerns included the potential for AI bias against certain populations, awareness of the resources used to develop AI tools, and broader questions about whether AI might eventually replace genetic counselors.

## 4. DISCUSSION

This study explores the impact of ambient AI documentation on genetic counselor burnout and examines its adoption considerations for clinical genetics workflows. Our mixed methods approach allowed us to quantitatively assess burnout patterns over time and contextualize them through qualitative analysis of interviews.

Reported use of ambient AI documentation was associated with short-term reduction in burnout. Across both our quantitative analyses methods (linear mixed-effects model and paired non-parametric Wilcoxon signed-rank test), ambient AI users demonstrated a decrease in burnout scores after 90 days followed by a modest increase at 270 days, though scores remained lower than pre-survey burnout scores. In contrast, for non-users, burnout scores changed minimally over time. Although this study was not powered to detect causal effects and the interaction between time and ambient AI use was not statistically significant, the consistency in the magnitude and direction of effects across our two statistical methods suggest that ambient AI use may reduce burnout in the short-term. These findings align with our qualitative findings and themes in existing literature that highlight reductions in cognitive and administrative burden of documentation.^2–3, 5, 36^

Interviews further highlighted which sections of genetic counseling notes were the most and least supported by ambient AI. Current ambient AI templates are designed for providers using SOAP templates, requiring counselors to customize these templates to include or emphasize genetic counseling note sections. Many participants lacked the time or technical confidence to make these customizations. This aligns with our quantitative findings: missing information or notes not being in the user’s style were the most important drivers for edits. Despite these challenges, most participants found the tool to be beneficial to summarize non-templated note elements central to clinical genetics visits, including family history, psychosocial counseling, complex decision-making, and descriptions of rare genetic conditions.

Ambient AI’s role as a memory aid also emerged as a clear finding in interviews, with our survey results and existing literature for other providers supporting this.^9^ At the 270-day timepoint, 100% of ambient AI users (n=7) agreed (somewhat agreed or strongly agreed) that ambient AI made them less worried about forgetting elements of a patient visit. Additionally, given that prior research has shown that a provider’s interaction with the EHR during visits can make patients feel as if the visit was impersonal or formulaic, our participants reported improvements in patient engagement by using more eye contact and better body language highlight a major benefit of this tool and align with prior findings.^5,10,11, 36, 37^

Participants had mixed opinions on accuracy of the generated notes. Attribution error occurred in group visits with multiple probands or family members. Findings in literature regarding this are mixed, with one study highlighting ambient AI struggling to distinguish between multiple speakers and another study stating it does not confuse speakers and functions well in distinguishing between different voices.^8, 38^

On a broader scale, our genetic counseling participants had similar concerns to other providers about data security and privacy, replacement of their role by AI, the need to balance optimization of healthcare efficiency using AI with empathy and compassion-focused care, and AI being a tool rather than a replacement.^37, 39–41^ These findings reinforce the need for implementation strategies that prioritize patient trust, transparency, and provider oversight.

When analyzing our qualitative data using the Consolidated Framework for Implementation Research (CFIR) framework, our findings align with key CFIR constructs influencing the implementation of ambient AI scribes for genetics providers.^34^ This tool’s relative advantage was in reducing documentation burden and improving patient engagement, while adaptability and complexity were barriers due to physician-centric templates and customization demands for genetic counseling notes. Compatibility varied by visit type (group visit with other providers vs genetic counseling-only visit), and this tool’s readiness for implementation was limited by time and technical savviness needed to tackle the learning curve and customization. Mapping these determinants to CFIR highlights actionable strategies. Future implementation efforts should address documentation needs unique to genetics providers to improve adaptability and sustainability, including the development of a genetics-specific note template through integration with IT support, and customization support through educational tipsheets and departmental presentations.^5,9,42^

Our study has several strengths including our sample comprising genetic counselors across multiple specialties, high survey response, methodological triangulation, inclusion of both users and non-users of this tool, overlap of participants who completed both surveys and interviews, and qualitative explanations for observed statistical patterns. Limitations include our study being conducted at a single institution using a single ambient AI tool, with convenience sampling and a small sample size, leading to limited generalizability of our findings.

Future studies on this topic can be conducted across multiple institutions comparing different ambient AI scribes, larger sample sizes, and objective documentation metrics.^10^ Studies can also assess patient perspectives on the use of AI tools in genetics visits.

## 5. CONCLUSION

As the demand for clinical genetics services continues to grow, strategic planning by healthcare systems to meet these needs can include the implementation of ambient AI documentation.^19^ Our data provides evidence that ambient AI technology has the potential to reduce documentation burden and short-term burnout among genetics providers. Future implementation efforts should address documentation needs unique to genetics providers. Addressing ethical and regulatory considerations will be essential for response integration of these tools for clinical genetics services.

## 6. ACKNOWLEDGEMENTS

This study was funded by the National Society of Genetic Counselors (NSGC) Special Interest Group (SIG) Research Grant Award of $500. The funder played no role in the study design, data collection, analysis and interpretation of data, or the writing of this manuscript.

Support for statistical analysis was provided by the Center for Social Science Innovation at the University of Iowa. The authors would also like to thank the study participants for their participation.

## 7. AUTHOR CONTRIBUTIONS

Conceptualization: AN, JM, CAC; Grant Submission: AN, CAC; Methodology: AN, JM, JVT, CAC; Data collection: AN, JM, CAC; Quantitative Data Analysis: AN, JM, CLS; Qualitative Data Analysis: AN, JVT; Writing (original draft): AN, Writing (review and editing): AN, JM, JVT, CLS, CAC; Supervision: JM, CAC. All authors read and approved the final manuscript.

## 8. FUNDING

Statistical analysis for this study was funded by the National Society of Genetic Counselors (NSGC) Special Interest Group (SIG) Research Grant Award of $500.

## 9. CONFLICTS OF INTEREST

AN presented the preliminary findings from this project as part of a panel at the National Society of Genetic Counselors (NSGC) Annual Conference in Seattle in November 2025 and received an honorarium for this presentation. All other authors have no financial or non-financial competing interests.

The ambient AI system evaluated in this study was used under a standard institutional license. The ambient AI vendor had no role in the study design, data collection, analysis, interpretation, or manuscript preparation.

## 10. DATA AVAILABILITY

The data that supports the findings of this study is not publicly available due to their containing information that could compromise study participant privacy. Further information is available from the corresponding author upon request.

## Notes

### Competing Interest Statement

First author Anjali Narain presented the preliminary findings from this project as part of a panel at the National Society of Genetic Counselors (NSGC) Annual Conference in Seattle in November 2025 and received an honorarium for this presentation. All other authors have no financial or non-financial competing interests.

### Author Declarations

The University of Iowa Institutional Review Board (IRB) determined that this project did not constitute human subjects research (IRB #202508020).

