## Supplemental Tables and Methods for "Ambient AI Documentation in Clinical Genetics: Perspectives on Implementation and Impact on Burnout"

**Supplemental Table 1: Demographics of participants**

|  | **Pre-ambient AI Survey N=16 (%)** | **Interview Participants N=14 (%)** |
| --- | --- | --- |
| **Sex** |  |  |
| F | 14 (87.5) | 13 (92.86) |
| M | 2 (12.5) | 1 (7.14) |
| **Age** |  |  |
| Under 25 | 0 (0) |  |
| 25-34 | 12 (75) |  |
| 35-44 | 3 (18.75) |  |
| 45-54 | 1 (6.25) |  |
| 55+ | 0 (0) |  |
| **Years worked at UI Health Care** |  |  |
| 0-5 years | 10 (62.5) |  |
| 6-10 years | 4 (25) |  |
| 11-15 years | 2 (12.5) |  |
| 15+ years | 0 (0) |  |
| **Genetic Counseling Specialty** |  |  |
| Neurology (Adult/Pediatric) | 3 | 3 |
| Internal Medicine (Cardiology/ Nephrology) | 2 | 2 |
| Women’s Health (Prenatal/ Reproductive Endocrinology | 4 | 3 |
| Oncology | 4 | 4 |
| Otolaryngology/Ophthalmology | 1 | 1 |
| Pediatric Medical Genetics | 3 | 2 |

*****The survey sample (N=16) and interview sample (N=14) were partially overlapping. One participant in the survey sample filled out the pre-survey but did not fill out the 90d and 270d surveys. Additionally, five participants had split positions between two specialties.

**Supplemental Table 2**: Response rates for pre-ambient AI survey, 90-day survey and 270-day survey

| **Type of Survey** | **Respondents (n)** | **Denominator** | **Response Rate** |
| --- | --- | --- | --- |
| Pre-ambient AI | 16 | 25 | 64% |
| 90-day survey | 13 | 16 | 81% |
| 270-day survey | 13 | 16 | 81% |
| 270-day survey from 90-day survey completion cohort | 11 | 13 | 85% |
| Completed all three surveys | 11 | 16 | 69% |

*Although the response rates for completion of 90-day and 270-day surveys are identical, the sample of participants who completed the 90-day and 270-day surveys (row 2 and row 3) is not identical. Two participants who initially completed only the pre-ambient AI survey later provided responses to the 90-day survey at the 270-day mark; since the 90-day and 270-day surveys were highly similar, these responses were counted as 270-day survey data. One participant’s 270-day response was not collected as it fell outside the data collection window. Another participant only filled out the pre-Ambient AI survey but did not fill out the 90d or 270d survey, so their response was not used for data analysis.

### **SUPPLEMENTAL METHODS**

Linear mixed-effects models are an extension of regular linear models that are used to analyze data in which the same units or people (i.e., survey respondents) are measured more than once at different points in time. Rather than treating every observation in the data as independent, linear mixed-effects models can account for the fact that measurements from the same person or unit are related. To account for this, a linear mixed-effects model was estimated with random intercepts for individuals to examine changes in burnout following access to ambient AI. Respondents were then followed up with at 90-days and 270-days, at which time burnout was re-measured and respondents reported whether they had used ambient AI since their previous contact. Ambient AI use was measured at each post-baseline survey wave (90 and 270 days) and varied within individuals over time. While the total number of respondents reporting ambient AI use was the same at both waves, some respondents changed their usage status between waves. Ambient AI use was treated as a time-varying covariate, allowing the model to capture within-person changes in burnout associated with AI use across survey waves. The resulting panel is unbalanced, with respondents contributing between two and three observations. Time was modeled as a continuous variable ‘Wave_post’ centered at the 90-day survey, the first wave at which ambient AI use was possible (coded 0 for the 90-day survey and 1 for the 270-day survey). It captures elapsed time since the first post-exposure measurement, allowing burnout trajectories to be modeled linearly after access to ambient AI. An interaction between time and ambient AI use, ‘Wave_post x Ambient AI use’ was included to assess whether the association between ambient AI use and burnout differs across post-baseline survey waves. A positive coefficient indicates that burnout increased more among users relative to non-users. A non-significant coefficient suggests similar post-exposure burnout trajectories across groups. This is a cross-level interaction (between-person X within-person). Models were estimated using maximum likelihood and retained all available observations.
